# Differences in Motivators, Barriers, and Incentives between Black and White Older Adults for Participation in Alzheimer’s Disease Biomarker Research

**DOI:** 10.1101/2022.09.10.22279803

**Authors:** Johanne Eliacin, Angelina J. Polsinelli, Francine Epperson, Sujuan Gao, Sarah Van Heiden, Glenda Westmoreland, Ralph Richards, Mollie Richards, Chris Campbell, Hugh Hendrie, Shannon L. Risacher, Andrew J. Saykin, Sophia Wang

## Abstract

**Introduction:** The study aimed to identify strategies to increase older Black adults’ participation in Alzheimer’s disease (AD) biomarker research studies.

**Methods:** 399 community-dwelling Black and White older adults (age ≥ 55) who had never participated in AD research completed a survey about their perceptions of AD research involving blood draw, MRI, and PET.

**Results:** Although most participants expressed interest in AD biomarker research (Black participants: 63.0%, White participants: 80.6%), Black participants were significantly more hesitant than White participants (28.9% vs 15.1%), were more concerned about study risks, (30.8% vs. 11.1%) and perceived multiple barriers to participating in brain scans. Lack of information was perceived as a barrier to participation across groups (45.8%) and return of study results was perceived as a participation incentive (78.9-85.7%) (*P*s < .05).

**Discussion:** Strategies to increase Black older adult participation in AD research may include disseminating additional study information and return of results.

**Declaration of Interest:** None of the investigators have a conflict of interest. JE receives support from VA IK HX002283, NIA P30AG072976, and NIA P30AG010133. AJP receives support from NIA (NIA U01 AG057195) and Alzheimer’s Association (LDRFP-21-818464). SW receives support from multiple NIA grants (K23AG062555, P30AG072976, P30AG010133, and R21AG074179) and the VA for clinical services. She receives book royalties from APPI and DSMB consultant fees (total less than $2000/year). AJS receives support from multiple NIH grants (P30 AG010133, P30 AG072976, R01 AG019771, R01 AG057739, U01 AG024904, R01 LM013463, R01 AG068193, T32 AG071444, and U01 AG068057 and U01 AG072177). He has also received support from Avid Radiopharmaceuticals, a subsidiary of Eli Lilly (in kind contribution of PET tracer precursor); Bayer Oncology (Scientific Advisory Board); Eisai (Scientific Advisory Board); Siemens Medical Solutions USA, Inc. (Dementia Advisory Board); Springer-Nature Publishing (Editorial Office Support as Editor-in-Chief, Brain Imaging and Behavior).

## INTRODUCTION

Although Black older adults are twice as likely as White older adults to develop Alzheimer’s disease and related dementias (ADRD) [1, 2], their participation rates in Alzheimer’s disease (AD) studies are significantly lower than White older adults’ [3–5]. Despite interest in AD research [6], they are often hesitant to enroll in intensive AD biomarker studies [7–10]. Addressing this hesitancy is essential to answering key questions about the underlying causes of health disparities in ADRD [11–13], particularly in the context of increasing focus on early detection with biomarkers and precision medicine.

Several studies have examined barriers to AD biomarker research participation in Black adults [7, 14–17]. However, many of these included subgroups of individuals who were already enrolled in AD research studies or registries, disproportionately female, highly educated, and had higher socioeconomic status. Historically, those with lower education and socioeconomic status have been underrepresented in AD research despite being at higher risk of developing the disease [18–20]. As researchers design community-based recruitment strategies to enroll new participants and broaden the pool of potential participants, one challenge is recruiting individuals from racially and ethnically diverse backgrounds who have never participated in AD research and have lower education and socioeconomic status [15].

To address these gaps, we examined interest, motivators, barriers, and incentives for participation in AD biomarker research for Black older adults who had never participated in AD research. We also focused on including subgroups with traditionally lower rates of research participation (i.e., men, < 16 years of education, lower socioeconomic [SES] status). Of note, this exploratory study was part of a larger research project – Promoting Cultural Awareness and Diversity in Research about Alzheimer’s Disease and Cognitive Health (AD-REACH) – which ultimately aims to generate targeted and culturally sensitive recruitment materials for Black older adults to increase recruitment into AD biomarker studies.

## METHODS

### 2.1 Study design and participants

This cross-sectional study involved survey data collected from community-dwelling older adults who met study eligibility criteria, including: 1) Identified as non-Hispanic White or African-American/Black (including biracial or multiracial), 2) age ≥ 55 years, 3) resided in the Indianapolis-Carmel-Anderson metropolitan area as defined by the Census Bureau Statistical Area map, and 4) had <u>never</u> participated in an Alzheimer’s disease research study.

We recruited participants through the Indiana University School of Medicine and the Roudebush VA Medical Center, from September 2021 to April 2022. Recruitment strategies included word-of-mouth from Indiana Alzheimer’s Disease Research Center (IADRC) study participants, IADRC staff, and IADRC Community Advisory Board (CAB) members; online advertisements; presentations at meetings with community-based partners; invitation mailings and phone calls made to potentially eligible participants identified via electronic health records at IU Health and Roudebush VA; and invitation emails sent to eligible participants in the ALLIN4HEALTH research registry (an Indiana University affiliated Clinical and Translational Sciences Institute sponsored research volunteer registry). Participants choose whether to complete the study survey over the phone with an IADRC staff member or independently online. The Indiana University Institutional Review Board (IRB) reviewed and approved the survey, and participants provided informed consent. Participants were compensated $35 for completing the survey. See **Supplementary Figure 1** for recruitment and eligibility diagram.

**Figure 1.**
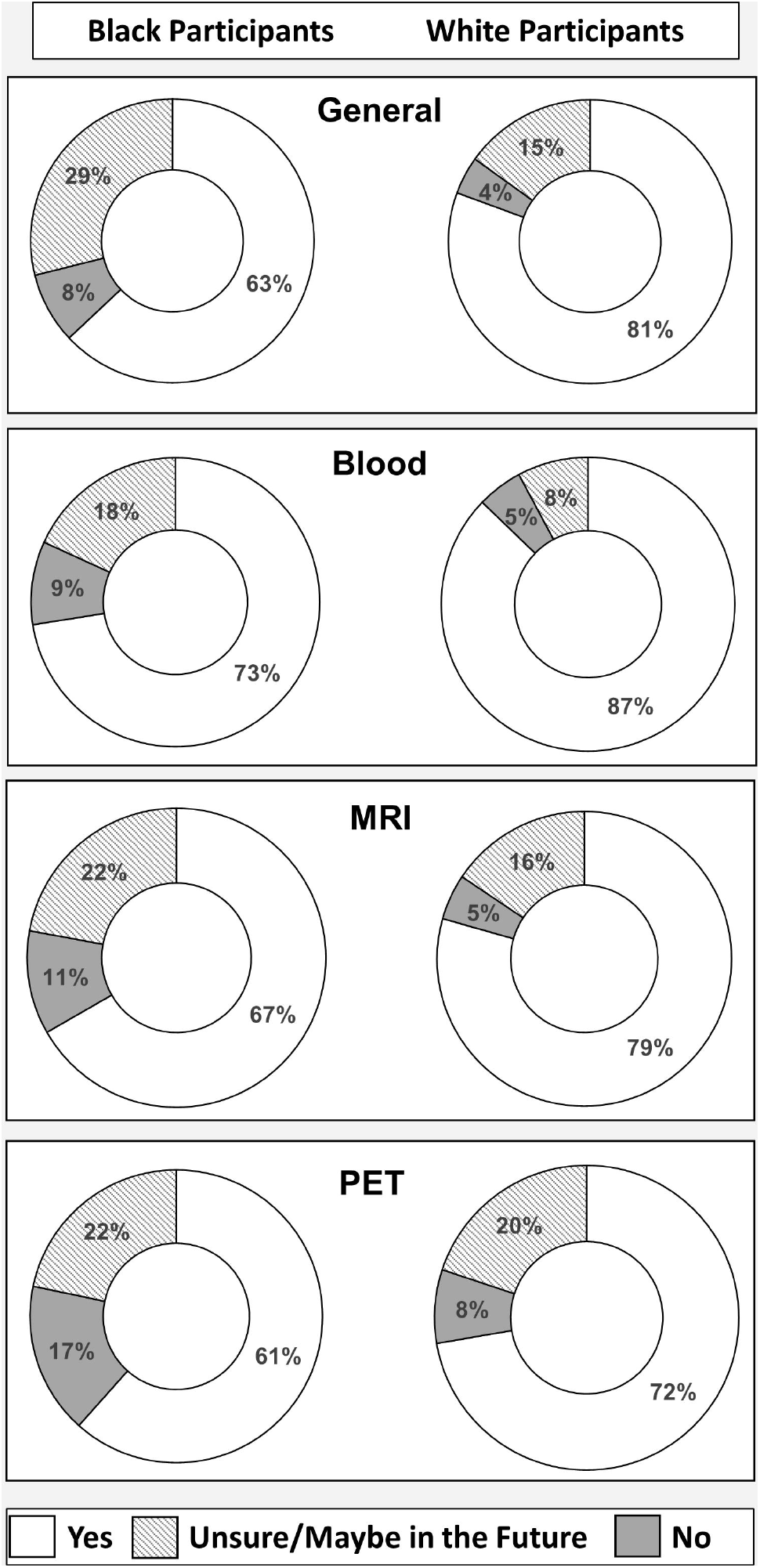
Percentage of participants interested (‘yes’), unsure (‘unsure’ or “maybe in the future’), and uninterested (‘no’) in participation in general Alzheimer’s disease biomarker research (n = 385), and in participation in specific biomarker procedures – blood draw for genetics, magnetic resonance imaging (MRI), or positron emission tomography (PET) (n = 399) – as a function of race.

### 2.3 Survey development

The AD-REACH survey was designed by conducting a series of focus groups and semi-structured qualitative interviews with racially diverse participants that included IADRC CAB members, IADRC study participants, and community-dwelling civilians and veterans [21]. Based on the themes identified from these interviews, we developed a preliminary version of the AD-REACH survey that we tested with a different sample of 10 individuals who met the same inclusion and exclusion criteria listed above. Study staff from the Center for Survey Research at IU Bloomington conducted cognitive interviewing [22] with these 10 individuals to assess their comprehension and perception of the survey, and provided our study team with suggestions for finalization of the survey.

### 2.4 Survey content

The AD-REACH survey covered several topics, including knowledge of AD, trust of researchers, health disparities of AD, and future interest in AD biomarker research participation. (See **Supplementary Figure 2** for the subset of AD-REACH survey questions and responses relevant to the present study.) For the present study, we focused on interest in future AD research participation and common biomarker procedures (blood sample for genetics, brain MRI scans, and PET scans), motivators and barriers to participating in AD biomarker research, and incentives for participation. Depending on participants’ interest in AD research – either general interest or specific procedural interest – we inquired about their motivations (those who expressed interest or hesitancy) and barriers (those who expressed lack of interest or hesitancy). (See **Supplementary Figures 3A-C** for flow diagrams of survey questions.) Finally, all participants were asked about incentives for participating in AD biomarker research including, brain health information, transportation/travel vouchers, and return of results for standard labs, cognitive testing, and neuroimaging. Gift cards and/or monetary incentives were not included in the list of incentives. Survey respondents were informed that these incentives would be in addition to gift cards.

**Figure 2.**
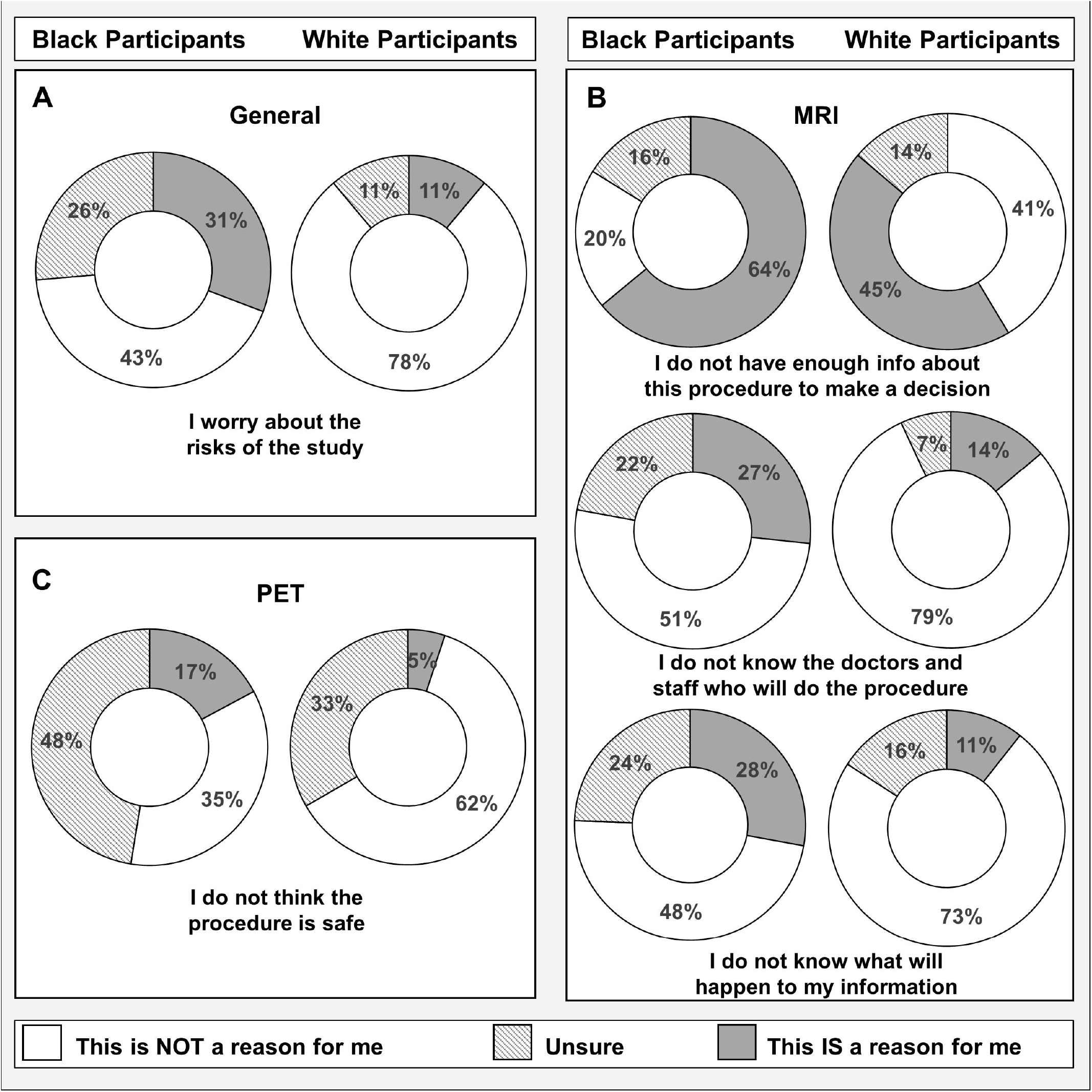
Percentage of Black and White participants endorsing (A) concern about safety of Alzheimer’s disease biomarker research in general; (B) barriers to magnetic resonance imaging (MRI) (n = 138) and; (C) concern about safety of positron emission tomography (PET) (n = 115).

**Figure 3.**
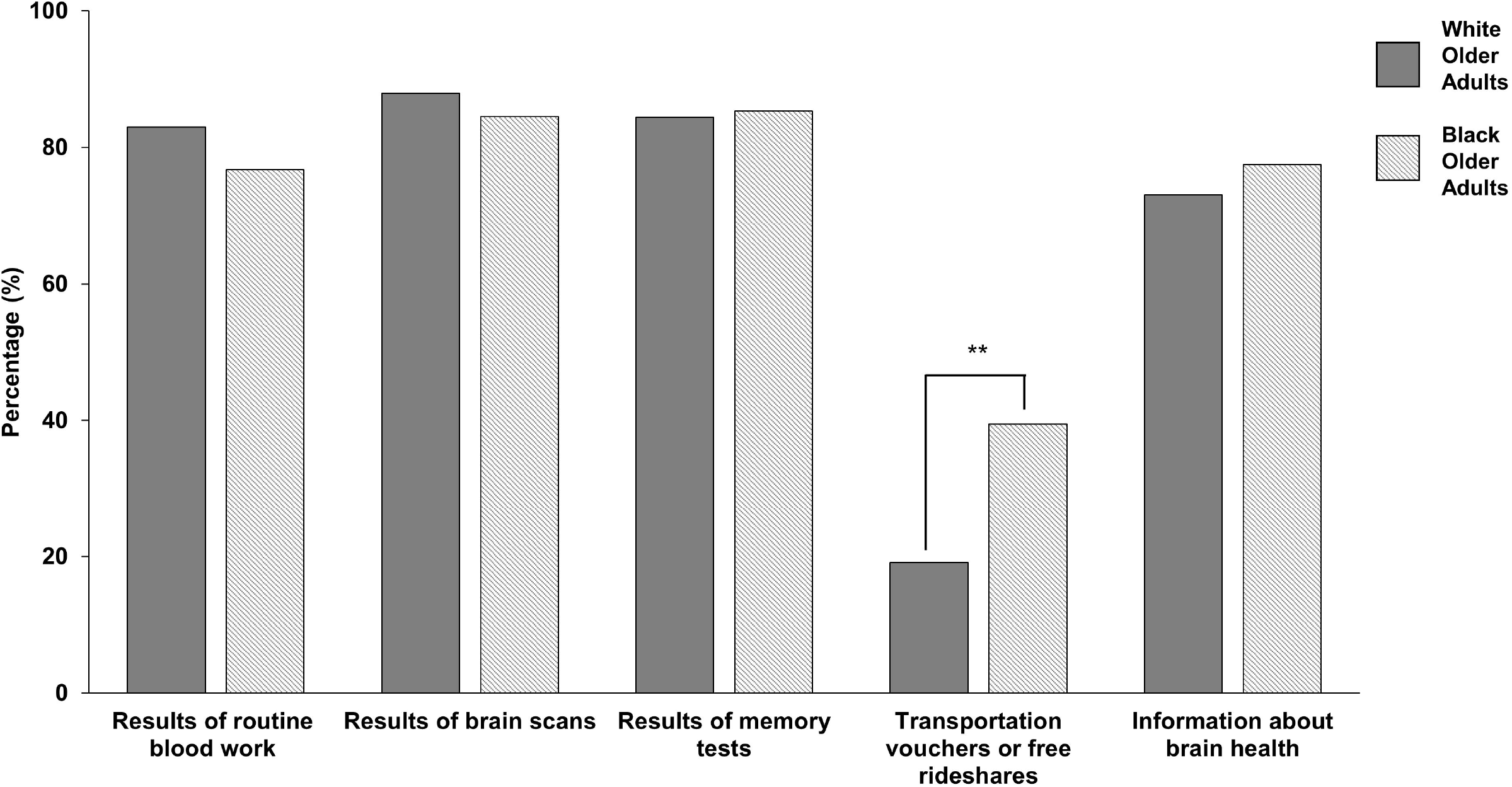
Percentage of Black and White participants endorsing each incentive for participating in Alzheimer’s disease biomarker research (n = 399).

### 2.5 Measures

#### 2.5.1. Demographic variables

‘Black participants’ is used to describe participants who self-identified as African American, Black, or biracial/multiracial. Age (55-64 years vs. > 65 years), education (< 16 years vs. >16 years), and sex (male vs. female) were dichotomized variables due to IRB regulations regarding deidentified survey collection. Estimated annual income (estimated using US Census Bureau data of median income for zip code) was categorized into quartiles.

#### 2.5.2. Knowledge of AD

Participants indicated their perceived knowledge of AD on one question with a 5-point scale. A score of 5 indicated highest level of knowledge while a score of 1 reflected no knowledge of AD. The mean score was 3.05 (SD = 0.80). We dichotomized this score based on the mean and whether participants reported knowing something about AD (*i.e*., higher level of knowledge of AD, ≥ 3) or not knowing anything about AD (*i.e*., lower level of knowledge of AD, < 3). Perceived knowledge of AD was included in our models (described below) as it varies as a function of race and can affect decisions to participate in AD research in underrepresented populations [23, 24].

#### 2.5.3 Trust of Researchers

Participants indicated their trust of researchers via six survey questions with three response choices – “agree” [score = 2], “unsure” [score = 1], “disagree” [score = 0]. The sum of these six questions reflected overall level of trust. Since the sum was skewed (mean = 11.2, SD = 1.8), we dichotomized this variable into those expressing trust on all questions (*i.e*., very high trust level, = 12) versus those who expressed hesitancy or lack of trust on any of the questions (*i.e*., low to high level of trust, ≤ 11). Trust of researchers was included in our models as it can be asignificant sociocultural factor affecting decision to participate in AD biomarker research [14, 25–27].

#### 2.5.4. Outcomes

Outcome variables were the responses to individual survey questions about AD biomarker research. These questions included interest in future participation in AD biomarker research generally and in specific procedures – blood work for genetics, MRI, and PET (‘yes’, ‘no’, or ‘unsure/maybe in the future’); motivators in participation (‘this is a reason for me’, ‘this is not a reason for me’, or ‘unsure’), barriers in participation (‘this is a reason for me’, ‘this is not a reason for me’, or ‘unsure’), and incentives for participating (select all responses that apply).

#### 2.5.5. Data analysis

We used chi-square tests to compare Black and White participants on demographics, knowledge of AD, and trust of researchers. We examined the association between race and responses to the survey questions using logistic regression models for each survey question. For outcome variables with only two options (“Yes” or “No”), we defined the reference group for the binary logistic regression model as “Yes.” For outcome variables with three response options (Yes/Unsure/No, Yes/Maybe in the future/No”, or This is a reason for me/Unsure/This is not a reason for me) we used ordinal logistic regression and defined Yes/This is not a reason for me = 0, Unsure/Maybe in the future = 1, and No/This is a reason for me = 2. Cumulative odds ratios (OR; ordinal) or OR (binary) and 95% confidence intervals (CIs) were estimated for each of logistic regression model.

In the ordinal and binary logistic regression models, we adjusted for covariates. In the first, demographic model, we adjusted for demographic variables (age, education, and sex). In the second, multivariate model, we adjusted for the same demographic variables and added knowledge about AD and trust of researchers. Due to the collinearity of education, marital/long-term partner status, and income, we included only education in the logistic regression models. The only exception was for the analysis of the transportation/travel vouchers incentive. For this model, we used estimated annual income instead of education because this was a monetary incentive.

Statistical tests were 2-tailed with *P* < .05 defining statistical significance. All analyses were performed using SPSS Statistics version 28.0. Given the exploratory nature of the study, we did not correct for multiple comparisons.

## RESULTS

### 3.1. Sample characteristics

258 Black and 141 White participants completed the AD-REACH survey. See **Table 1** for participants’ demographics. Black participants were more likely than White participants to have less than 16 years of education, less likely to be married or have a lifetime partner and more likely to be in the lowest quartile of estimated annual income (*P*s < .05). There were no statistically significant race differences for sex or age.

**Table 1.** Participant demographics

|  | Black older adults<br>n = 258<br>% (n) | White older adults<br>n = 141<br>% (n) |
| --- | --- | --- |
| **Age (years)** |  |  |
| 55-64 | 42.2 (109) | 34.8 (49) |
| >65 | 57.8 (149) | 65.2 (92) |
| **Sex** |  |  |
| Female | 53.5 (138) | 61.7 (87) |
| Male | 42.5 (120) | 38.3 (54) |
| **Education** |  |  |
| <16 years | 73.3 (189)* | 41.1 (58)* |
| >16 years | 26.7 (69)* | 58.9 (83)* |
| **Relationship** |  |  |
| Married/partnered | 42.6 (110)* | 64.5 (91)* |
| Single | 57.4 (148)* | 35.5 (50)* |
| **Estimated annual income level** |  |  |
| 1<sup>st</sup> quartile (< 40,707) | 29.1 (75)* | 11.3 (16)* |
| 2<sup>nd</sup> quartile (40,707-52,068) | 26.7 (69) | 26.2 (37) |
| 3<sup>rd</sup> quartile (52,068-72,928) | 24.8 (64) | 22.7 (32) |
| 4<sup>th</sup> quartile (> 72,928) | 19.4 (50)* | 39.7 (56)* |
| **Knowledge of AD** |  |  |
| ≤ 2 | 24.0 (62)* | 5.0 (7)* |
| ≥ 3 | 76.0 (196)* | 95.0 (134)* |
| **Trust of Researchers** |  |  |
| ≤ 11 | 34.5 (89)* | 19.1 (27)* |
| 12 | 65.5 (169)* | 80.9 (114)* |

NOTE: \* $p < .05$ as per chi-squared tests. Annual income levels were estimated using the US Bureau Census data for the median income for the reported zip codes.

### 3.2 Trust of researchers and knowledge of AD

Black participants were less likely than White participants to report that they knew about AD and had very high trust of researchers (*P*s ≤ .001). Nearly all participants (96.5%, *N* = 385) indicated that they had never been invited to participate in an AD research study. Reasons for not participating in AD research for the 14 participants who had previously been invited are described in **Supplementary Figure 4**.

### 3.3 Main analyses

### 3.3.1. Interest in participation in a future AD biomarker study and specific AD biomarker procedures

**Table 2A** presents the ORs of the logistic regression models comparing Black and White participants’ hesitancy in AD biomarker research. Most participants, regardless of race, expressed interest in general AD biomarker research (Black participants: 63.0%; White participants: 80.6%). However, Black participants were overall more hesitant than White participants (multivariate OR = 1.75, 95% CI 1.01-3.04, *P* = .045). Black participants were also more hesitant than White participants about all specific AD biomarker procedures, including blood draw for genetics (demographic OR = 2.24, 95% CI 1.24-4.04, *P* = .008), brain MRI (demographic OR = 1.78, 95% CI 1.07-2.96, *P* = .027), and PET scan (demographic OR = 1.78, 95% CI 1.11-2.84, *P* = .017). However, there were no significant race differences after adjusting for knowledge about AD and trust of researchers (*P*s > .05). See **Figure 1** for the percentage of responses reflecting interest (‘yes’), uncertainty (‘unsure/maybe in the future’), and disinterest (‘no”) in AD biomarker research.

**Table 2.** (A) Odds ratios and 95% confidence intervals for differences in Black vs. White participants’ hesitancy in Alzheimer’s disease (AD) biomarker research, generally, and related to specific biomarker procedures; (B) Odds ratios and 95% confidence intervals for differences in Black vs. White participants’ selections of incentives to participate in AD biomarker research

| A. | demographic adjusted |  |  |  | multivariate adjusted |  |  |  |
| --- | --- | --- | --- | --- | --- | --- | --- | --- |
|  | 95% CI |  |  |  | 95% CI |  |  |  |
|  | OR | lower | upper | p | OR | lower | upper | p |
| General (n = 385) |  |  |  |  |  |  |  |  |
| Would you consider AD research? | 2.40 | 1.42 | 4.03 | **0.001** | 1.75 | 1.01 | 3.04 | **0.045** |
| Specific (n = 399) |  |  |  |  |  |  |  |  |
| Would you consider blood draw? | 2.24 | 1.24 | 4.04 | **0.008** | 1.70 | 0.92 | 3.16 | 0.090 |
| Would you consider MRI? | 1.78 | 1.07 | 2.96 | **0.027** | 1.59 | 0.94 | 2.69 | 0.084 |
| Would you consider PET? | 1.78 | 1.11 | 2.84 | **0.017** | 1.56 | 0.96 | 2.54 | 0.073 |
| B. | demographic adjusted |  |  |  | multivariate adjusted |  |  |  |
|  | 95% CI |  |  |  | 95% CI |  |  |  |
|  | OR | lower | upper | p | OR | lower | upper | p |
| (n = 399) |  |  |  |  |  |  |  |  |
| results of routine blood work | 0.74 | 0.42 | 1.30 | 0.300 | 0.90 | 0.50 | 1.62 | 0.729 |
| results of brain scans | 0.85 | 0.45 | 1.63 | 0.631 | 1.13 | 0.57 | 2.23 | 0.730 |
| results of memory tests | 1.08 | 0.60 | 1.99 | 0.800 | 1.36 | 0.72 | 2.59 | 0.340 |
| transportation vouchers | 2.39 | 0.44 | 3.96 | **<.001** | 2.29 | 1.36 | 3.86 | **0.002** |
| information about brain health | 1.30 | 0.78 | 2.15 | 0.312 | 1.32 | 0.78 | 2.22 | 0.302 |
| return of normal results | 1.06 | 0.49 | 2.40 | 0.900 | 0.84 | 0.35 | 1.98 | 0.690 |

NOTE: OR = odds ration; CI = confidence interval. Cumulative OR and 95% CI were calculated using ordinal logistic regression models adjusting for only demographic variables (sex, age, education; "demographic adjusted") and demographic variables in addition to knowledge of AD and trust of researchers ("multivariate adjusted").

### 3.3.2. Barriers to participation in AD biomarker research and specific AD biomarker procedures

**Table 3** presents the ORs of the logistic regression models comparing Black and White participants’ perceived barriers to AD research. The two general AD biomarker research barriers most frequently endorsed by both Black and White participants were: “I would consider participating in a study now, but I need more information” and “I do not know enough about Alzheimer’s disease to make a decision about participating in a research study” (*P*s > .05). Black participants were more concerned about general AD biomarker study risks than White participants (multivariate OR = 3.2, 95% CI 1.13-9.53, *P* = .029) (**Figure 2A**). Examining individual procedures, the two most frequently endorsed barriers were the same for all procedures (blood draw, MRI, PET): “I do not have enough information about this procedure to make a decision” and “I only do these procedures when they are medically necessary”. Black participants identified multiple barriers to participating in brain scans compared to White participants. For MRI, these included, not having enough information to decide about participating (multivariate OR = 2.59, 95% CI 1.04-6.44, *P* = .041), not knowing the doctors/staff who would perform the procedure (multivariate OR = 3.03, 95% CI 1.06-8.63, *P* = .038), and not knowing what would happen to their information (multivariate OR = 3.55, 95% CI 1.07-10.44, *P* = .037) (**Figure 2B**). For PET, Black participants expressed greater concern than White participants about the safety of the procedure (multivariate OR = 2.93, 95% CI 1.27-6.77, *P* = .012) (**Figure 2C**). See **Supplementary Figures 5A-D** for the full data on perceived barriers.

**Table 3.** Odds ratios and 95% confidence intervals for differences in Black vs. White participants’ reasons for participating and NOT participating in Alzheimer’s disease (AD) biomarker research, generally, and related to specific biomarker procedures.

|  | demographic adjusted |  |  |  | multivariate adjusted |  |  |  |
| --- | --- | --- | --- | --- | --- | --- | --- | --- |
|  | 95% CI |  |  |  | 95% CI |  |  |  |
|  | OR | lower | upper | p | OR | lower | upper | p |
| **Reasons to participate** |  |  |  |  |  |  |  |  |
| *General (n = 385)* |  |  |  |  |  |  |  |  |
| loved one or I could be diagnosed | 0.86 | 0.42 | 1.76 | 0.68 | 0.96 | 0.46 | 2.028 | 0.92 |
| help find a cure | 1.89 | 0.88 | 4.02 | 0.10 | 2.21 | 0.99 | 4.914 | 0.05 |
| learn more about AD | 0.40 | 0.14 | 1.12 | 0.08 | 0.51 | 0.17 | 1.47 | 0.21 |
| everyone should do research | 0.76 | 0.49 | 1.19 | 0.23 | 0.89 | 0.56 | 1.415 | 0.61 |
| **Reasons NOT to participate** |  |  |  |  |  |  |  |  |
| *General (n = 118)* |  |  |  |  |  |  |  |  |
| do not know enough to decide | 0.99 | 0.39 | 2.49 | 0.98 | 0.85 | 0.33 | 2.23 | 0.75 |
| it will not make a difference | 1.62 | 0.47 | 5.60 | 0.45 | 1.40 | 0.39 | 5.01 | 0.34 |
| worry how information will be used | 1.32 | 0.48 | 3.65 | 0.59 | 0.97 | 0.33 | 2.84 | 0.95 |
| worry about abnormal results | 0.85 | 0.32 | 2.28 | 0.75 | 0.90 | 0.33 | 2.51 | 0.85 |
| worry about risks | 3.46 | 1.23 | 9.70 | **0.02** | 3.27 | 1.13 | 9.53 | **0.03** |
| not interested in AD research | 1.03 | 0.33 | 3.21 | 0.95 | 1.17 | 0.37 | 3.10 | 0.79 |
| need more information | 1.45 | 0.60 | 3.49 | 0.41 | 1.26 | 0.51 | 3.14 | 0.64 |
| *Blood (n = 89)* |  |  |  |  |  |  |  |  |
| do not have enough info to decide | 2.57 | 0.89 | 7.45 | 0.08 | 2.87 | 0.96 | 8.53 | 0.06 |
| worry procedure is unsafe | 1.06 | 0.35 | 3.16 | 0.92 | 1.03 | 0.34 | 3.11 | 0.96 |
| do not know the people doing the procedure | 0.91 | 0.30 | 2.78 | 0.87 | 1.03 | 0.33 | 3.17 | 0.96 |
| concern about what happens with information | 2.54 | 0.80 | 8.07 | 0.11 | 3.16 | 0.96 | 10.38 | 0.06 |
| only do this when medically necessary | 0.85 | 0.29 | 2.53 | 0.77 | 0.86 | 0.29 | 2.57 | 0.78 |
| worry about abnormal results | 0.94 | 0.31 | 2.82 | 0.91 | 0.92 | 0.30 | 2.84 | 0.89 |
| *MRI (n = 115)* |  |  |  |  |  |  |  |  |
| do not have enough info to decide | 2.92 | 1.20 | 7.13 | **0.02** | 2.59 | 1.04 | 6.44 | **0.04** |
| worry procedure is unsafe | 1.66 | 0.63 | 4.35 | 0.30 | 1.40 | 0.52 | 3.83 | 0.51 |
| do not know the people doing the procedure | 2.95 | 1.06 | 8.26 | **0.04** | 3.03 | 1.06 | 8.63 | **0.04** |
| concern about what happens with information | 4.41 | 1.47 | 13.24 | **0.01** | 3.35 | 1.07 | 10.44 | **0.04** |
| only do this when medically necessary | 1.98 | 0.81 | 4.82 | 0.13 | 1.89 | 0.76 | 4.71 | 0.17 |
| worry about abnormal results | 1.40 | 0.56 | 3.49 | 0.47 | 1.06 | 0.40 | 2.77 | 0.91 |
| *PET (n = 138)* |  |  |  |  |  |  |  |  |
| do not have enough info to decide | 1.47 | 0.66 | 3.31 | 0.35 | 1.26 | 0.55 | 2.90 | 0.58 |
| worry procedure is unsafe | 3.31 | 1.47 | 7.46 | **0.00** | 2.93 | 1.27 | 6.77 | **0.01** |
| do not know the people doing the procedure | 2.02 | 0.89 | 4.57 | 0.09 | 2.10 | 0.91 | 4.85 | 0.08 |
| concern about what happens with information | 2.30 | 0.95 | 5.57 | 0.07 | 1.94 | 0.78 | 4.86 | 0.16 |
| only do this when medically necessary | 1.33 | 0.61 | 2.88 | 0.47 | 1.32 | 0.60 | 2.92 | 0.49 |
| worry about abnormal results | 1.50 | 0.67 | 3.37 | 0.32 | 1.34 | 0.59 | 3.09 | 0.49 |

NOTE: OR = odds ratio; CI = confidence interval. Cumulative OR and 95% CI were calculated using ordinal logistic regression models adjusting for only demographic variables (sex, age, education; "demographic adjusted") and demographic variables in addition to knowledge of AD and trust of researchers ("multivariate adjusted").

### 3.3.3. Motivators for participation in AD biomarker research

**Table 3** presents the ORs of the logistic regression models comparing Black and White participants’ motivators for participating in AD biomarker research. Nearly all Black and White participants endorsed the same motivators for research participation at similar rates. There were no race differences for any of these motivators (*P*s > .05). See **Supplementary Figure 6** for full data of motivators.

### 3.3.4. Incentives to increase participation in AD biomarker research

**Table 2B** presents the ORs of the logistic regression models comparing Black and White participants’ incentives for AD biomarker research. As shown in **Figure 3**, most participants, regardless of race, were interested in receiving results of cognitive testing (Black participants: 85.3%; White participants: 84.4%), brain scans (Black participants: 84.5%; White participants: 87.9%), standard labs (Black participants: 76.7%; White participants: 83.0%), and information about brain health (Black participants: 77.5%; White participants: 73.0%) (*P*s > .05). Both Black and White participants agreed that even normal results from the AD biomarker procedures should be shared (91.1% vs 92%) (*P* > .05). Black participants were more likely than White participants to be interested in transportation or travel vouchers (39.5% vs 19.1%; multivariate OR = 2.32, 95% CI 1.38-3.91; *P* = .002).

In a post-hoc analysis, we examined associations between incentives (and interest in participating in AD biomarker research (**Supplementary Table 1**). Return of results for all types of procedures was consistently associated with a 3- to 4- fold higher interest (all *P*s < .05). Information about brain health was associated with a 2-fold higher interest. Offering transportation was not significantly associated with interest levels (*P* = .12).

## DISCUSSION

In the present study, we examined motivators, barriers, and incentives for participation in AD biomarker research in a community sample of Black and White older adults with no history of AD research experience. Although most Black and White participants were interested in AD biomarker research, a larger proportion of Black participants expressed hesitancy. Perceived barriers were mostly similar between groups, particularly about having enough information to make a decision about participation. However, lack of information was a particular concern for Black older adults regarding brain scans (MRI and PET). Black and White older adults expressed similar reasons for being motivated to participate in AD biomarker research and desired similar informational incentives, particularly return of individual procedure results. Overall, these results suggest Black and White older adults share similar concerns and motivations for participating in AD biomarker research, but Black older adults remain more hesitant. Our results add to previous studies showing education about AD research, addressing barriers to participation in neuroimaging and biomarker research procedures, and providing appropriate incentives may facilitate participant interest in AD biomarker studies [5–8].

Several factors may account for differences in hesitancy towards AD biomarker research among Black and White participants. These include differences in trust of researchers and research practices as well as perceived knowledge of AD [14, 28, 29]. However, after accounting for these factors, Black participants were still overall more hesitant than White participants. This suggests there are likely additional factors at play (e.g., beliefs about personal risk of AD, research attitudes, perceived risk and safety) or interactions between various factors that are not accounted for in these race differences. Of note, the average level of trust of researchers in our groups was high. This likely reflects our sampling method (see limitations below) and could have reduced our ability to find a stronger association between trust and hesitancy. Regardless, there is a need for further work to examine why Black older adults consistently expressed hesitancy for AD biomarker studies. One possible hypothesis may be that Black older adults may be more risk averse when making decisions [7], particularly with incomplete information or uncertainty about the source of information.

A predominant theme from our study was that perceived lack of information was a barrier to participating in AD biomarkers studies, and this was particularly true for Black older adults. The most consistently endorsed barriers for both Black and White older adults were lack of knowledge of AD and AD research procedures. Black older adults also perceived more barriers to undergoing brain scans than White older adults, especially not knowing the health professionals involved in MRI or what would be done with MRI information, and concern that PET was unsafe. These findings corroborate previous work highlighting the importance of appropriate information dissemination [19] and suggest that greater education about AD and research procedures is necessary. During the process of recruiting both Black and White older adults, researchers need to explain to participants why these procedures are being conducted, what the procedure entails in appropriate detail, and provide opportunities to discuss safety concerns and risks. While education, information, and outreach appear to be necessary for successful recruitment strategies [3, 30, 31], participants also identified return of results as an important incentive for participation.

Providing information about AD, brain health, and personal health results were notable incentives for participation for both Black and White older adults. Although Black older adults’ perception of their AD knowledge was lower than White participants, they were motivated to participate in AD biomarker research to improve their understanding of the disease and ways to keep their brain healthy. Additionally, an overwhelming majority of Black and White older adults viewed return of results for study procedures, including cognitive testing and brain scans, as very strong incentives for AD biomarker research participation. Erickson et al [17] found similar results among individuals enrolled in AD research. Willingness to participate in biomarker procedures was higher if personal results were disclosed versus not disclosed. This effect was particularly pronounced for Black adults. In our study, there was also near universal agreement that participants should receive normal tests results. However, providing study results as an incentive to diversify AD biomarker participation is not a straightforward process [15]. About one-third of our participants who stated they were already hesitant about research indicated they were worried or uncertain about being given abnormal test results. This aligns with researchers’ concerns about causing psychological harm or creating uncertainty about what the meaning of results – particularly with asymptomatic individuals [15, 16, 32–35]. Future research is needed to determine how to ethically and meaningfully return results to participants, particularly those from diverse populations.

Black older adults also identified transportation assistance as a key incentive for research participation, and this was not solely driven by SES. Covering costs of transit or assisting in arranging transit minimizes one of the higher-burden barriers to research participation [15, 36, 37]. These results suggest that assisting with transportation costs may be important to this group, not only from a socioeconomic standpoint, but also in demonstrating efforts to relieve burden and increase equity and reciprocity between researchers and participants.

Major strengths of our study include the size and the diversity of participants who have been traditionally underrepresented in AD biomarker research, including Black men, those with less than a college education, and those of lower socioeconomic status are highly represented in this study. Similarly, the perspectives captured in this study are from individuals who have never participated in AD biomarker research. Our findings, therefore, have implications for new recruitment strategies for underrepresented populations and individuals who are relatively at higher risk for AD.

However, the present study is not without limitations. First, we restricted our survey population to Black older adults residing in the Indianapolis metropolitan area. Its findings therefore are not generalizable. Future studies should include urban and rural populations of different geographical areas, to reflect different sociocultural and geographical contexts that may influence views of AD research participation. Second, results likely reflect the perspectives of individuals who were highly motivated to complete the survey (i.e., selection bias) as we did not entirely rely on random sampling. Third, although the survey content was based on input from individuals of the target population, these data are constrained by pre-defined content and responses. Finally, as this was an exploratory study, we did not correct for multiple comparisons. Future studies will need to replicate these findings and may require mixed methods to more deeply understand the barriers, motivators, and decision-making processes of minoritized groups [8].

In summary, our study identified several promising recruitment strategies to increase Black older adult participation, including improving information sharing and communication about AD biomarker procedures, particularly brain scans, return of results from study procedures, and providing transportation assistance. Future studies will need to examine how to develop and appropriately implement these evidence-based, socioculturally sensitive recruitment strategies to successfully increase enrollment of Black older adults into AD biomarker research studies.

## Supporting information

List of supplemental Figures

## Data Availability

All data produced in the present study are available upon reasonable request to the authors

## Acknowledgements

We thank all the AD-REACH participants, the community-based organizations and their leadership who spread the word about the AD-REACH survey, Community Advisory Board members (notably Denise Harrington, Lynda Montgomery, Shokrina Beering, Hank Mosley, and Michelle Bellamy), Andrew Tackett, Alan Beard, Caprice Elliott, Sam Davies, and Donna Wert for their assistance with participant recruitment, data collection, data organization, and manuscript formatting assistance.

## Funding and Support

This research was supported by the National Institutes of Health’s National Institute on Aging, P30AG10133-30 (A.J.S., S.W.), P30 AG072976-01 (A.J.S., S.W., J.E.), and R01 AG019771 (A.J.S.) and by the Alzheimer’s Association, LDRFP-21-818464 (S.W., A.J.P.). This content is solely the responsibility of the authors and does not necessarily represent the official views of the National Institutes of Health’s National Institute on Aging or the Alzheimer’s Association.

This material was supported with resources and the use of facilities at the Roudebush VAMC. This research also does not represent the official view of the VA Medical Center. The contents do not represent the views of VA or the United States Government.

