## Supplementary figures and images for "Differences in Motivators, Barriers, and Incentives between Black and White Older Adults for Participation in Alzheimer’s Disease Biomarker Research"

### Supplemental Figure 1

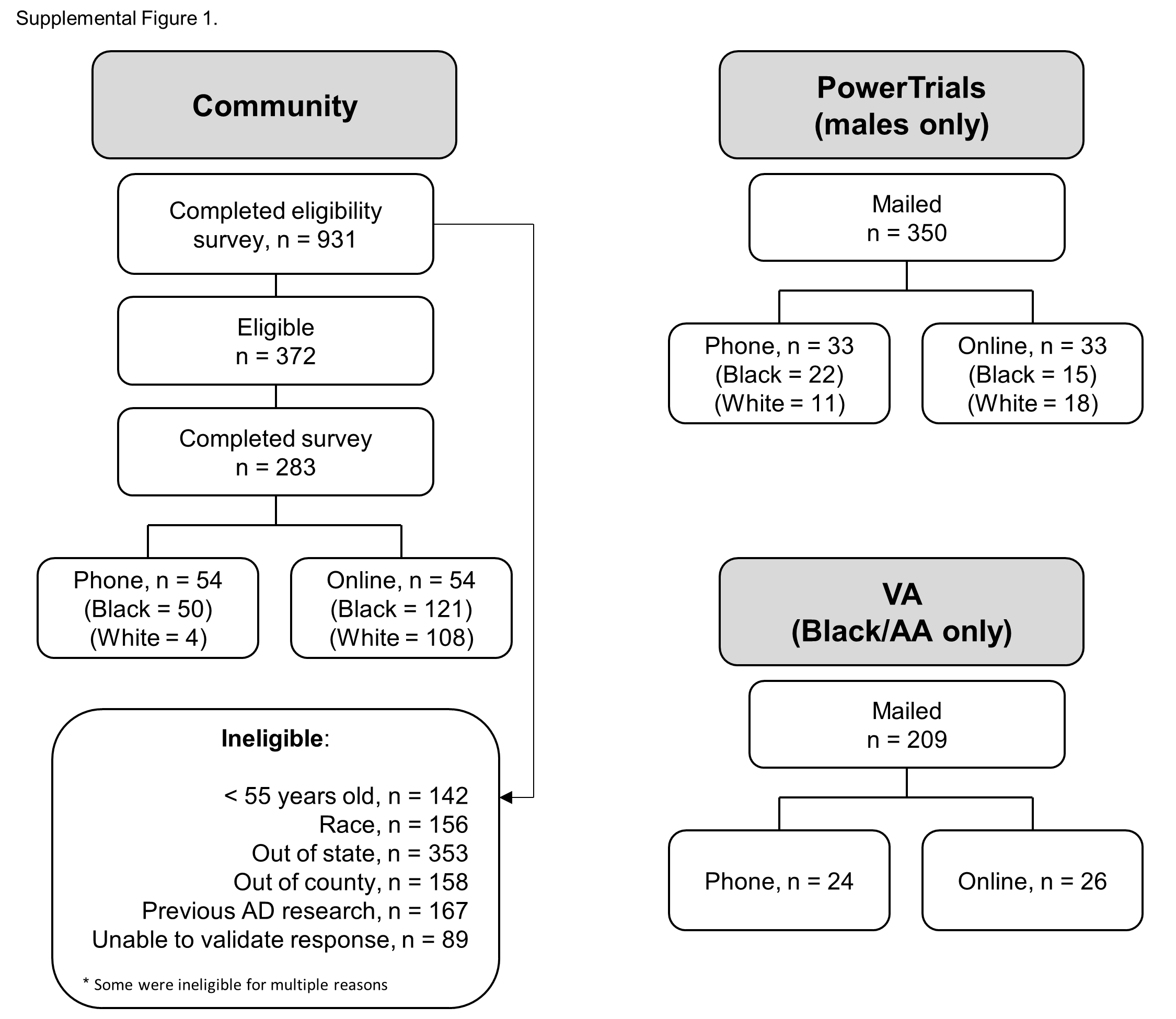

### Supplemental Figure 3

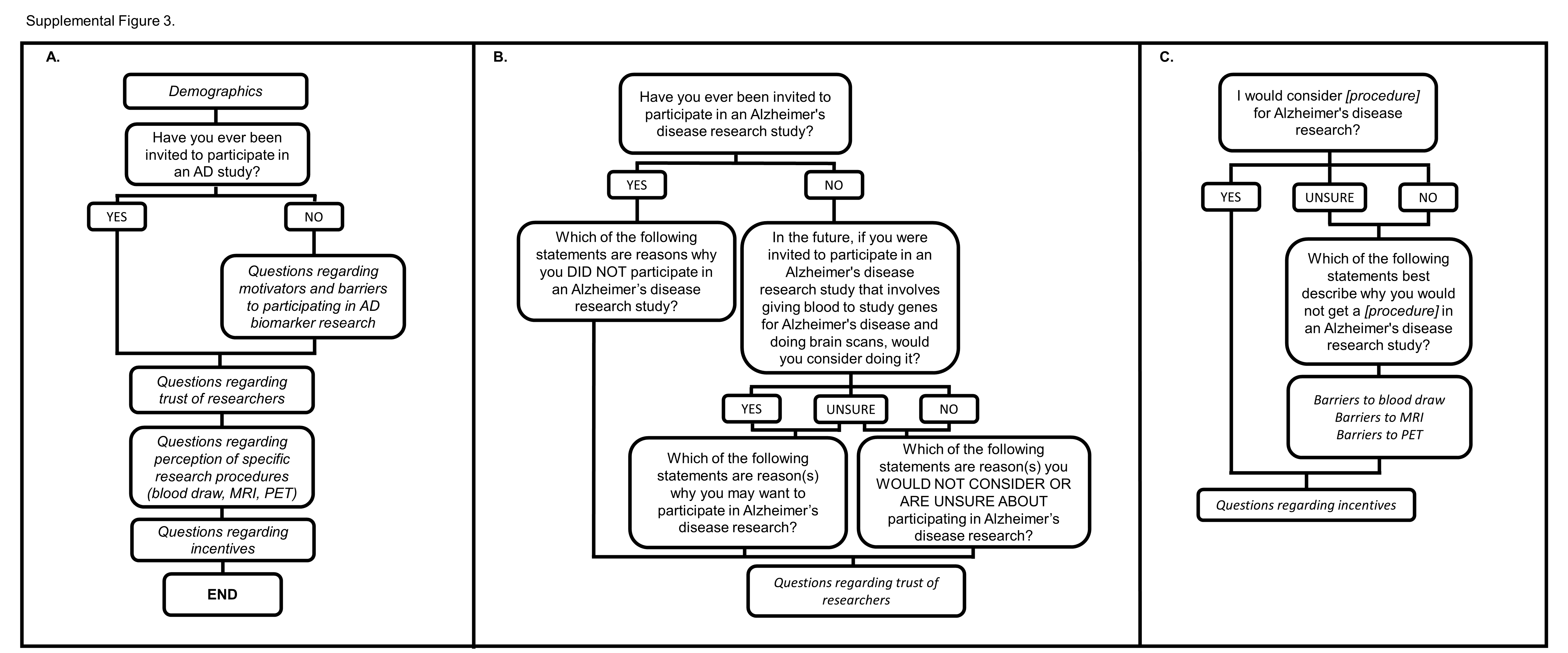

### Supplemental Figure 4

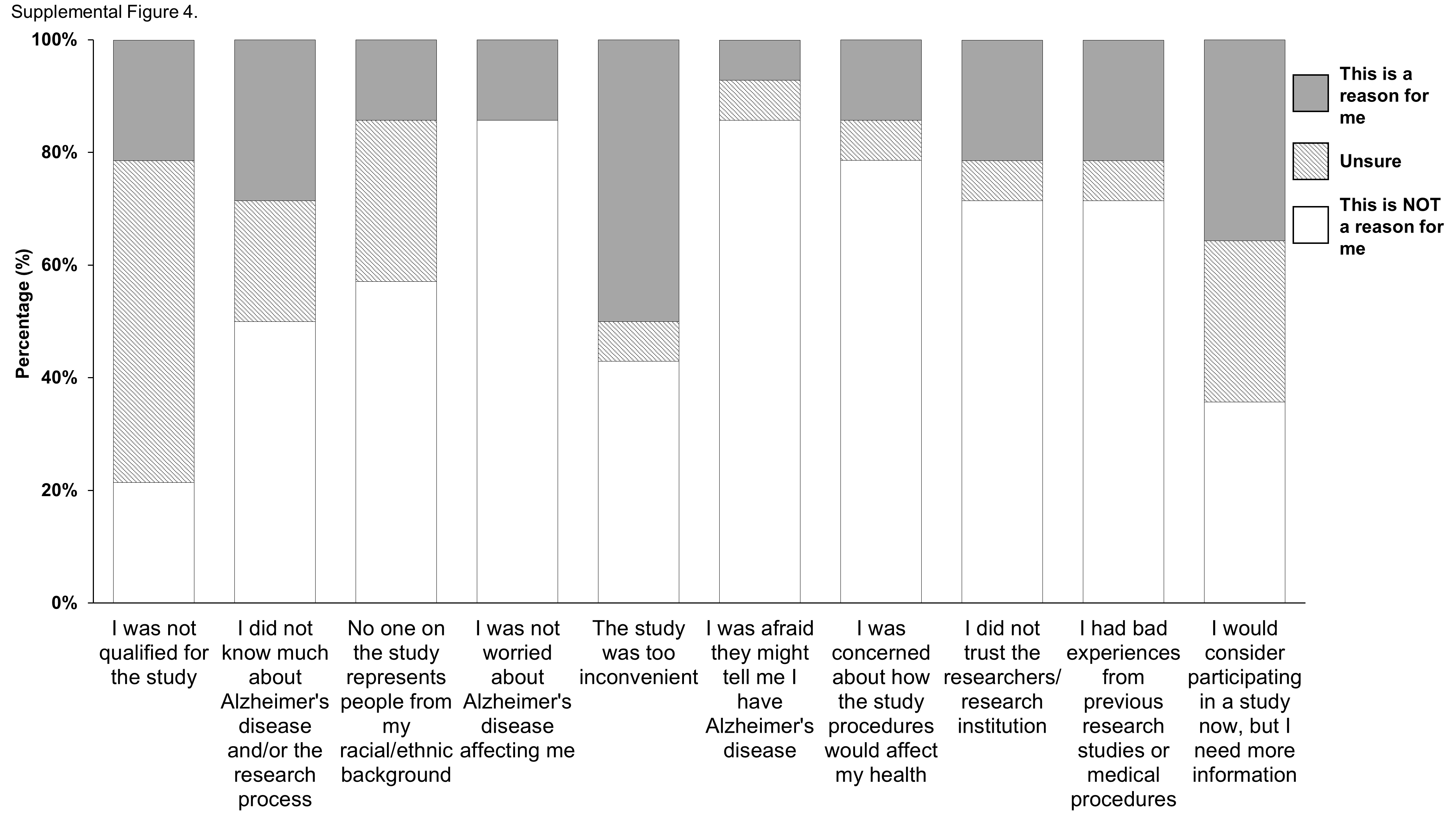

### Supplemental Figure 5

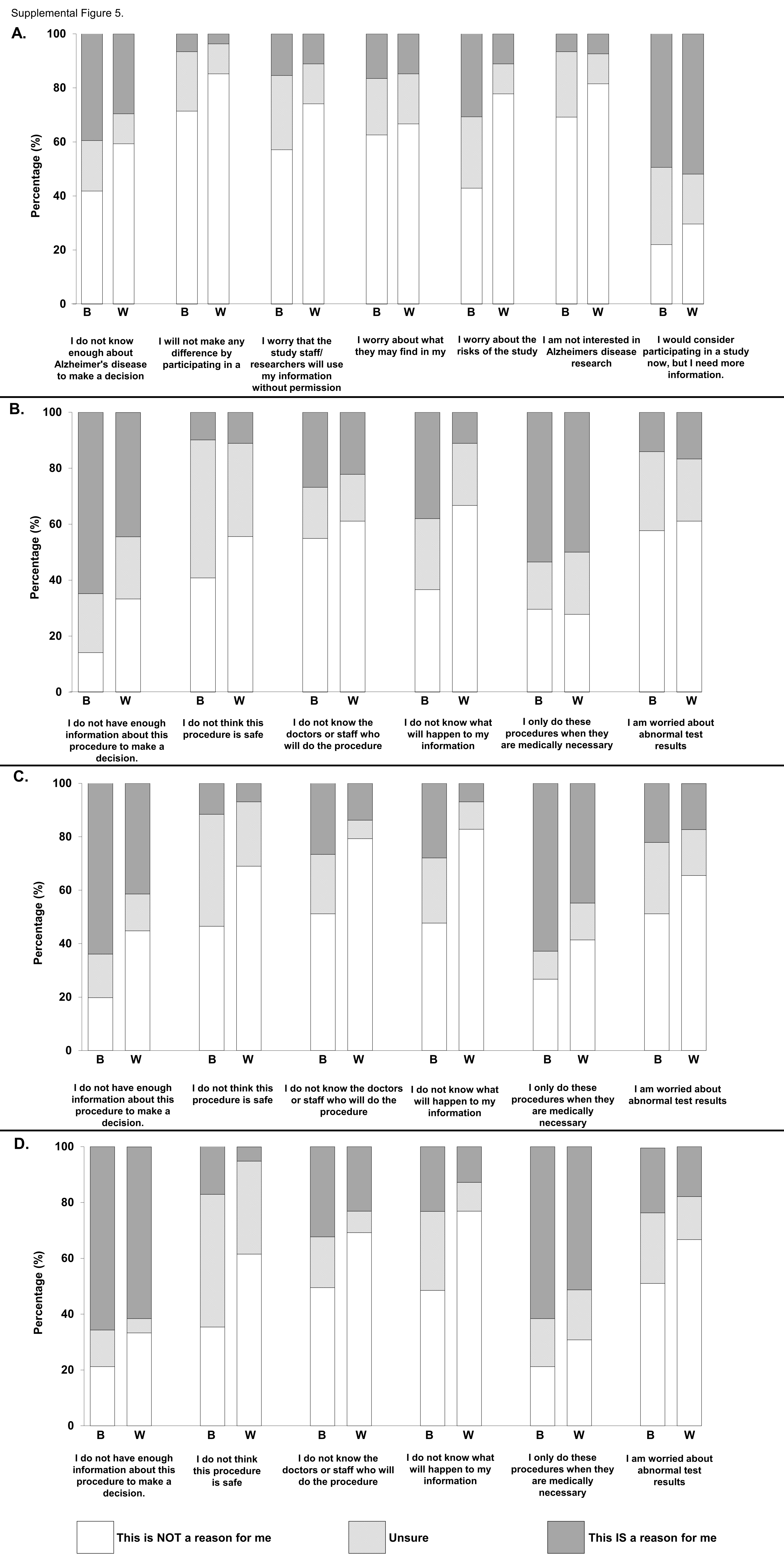

### Supplemental Figure 6

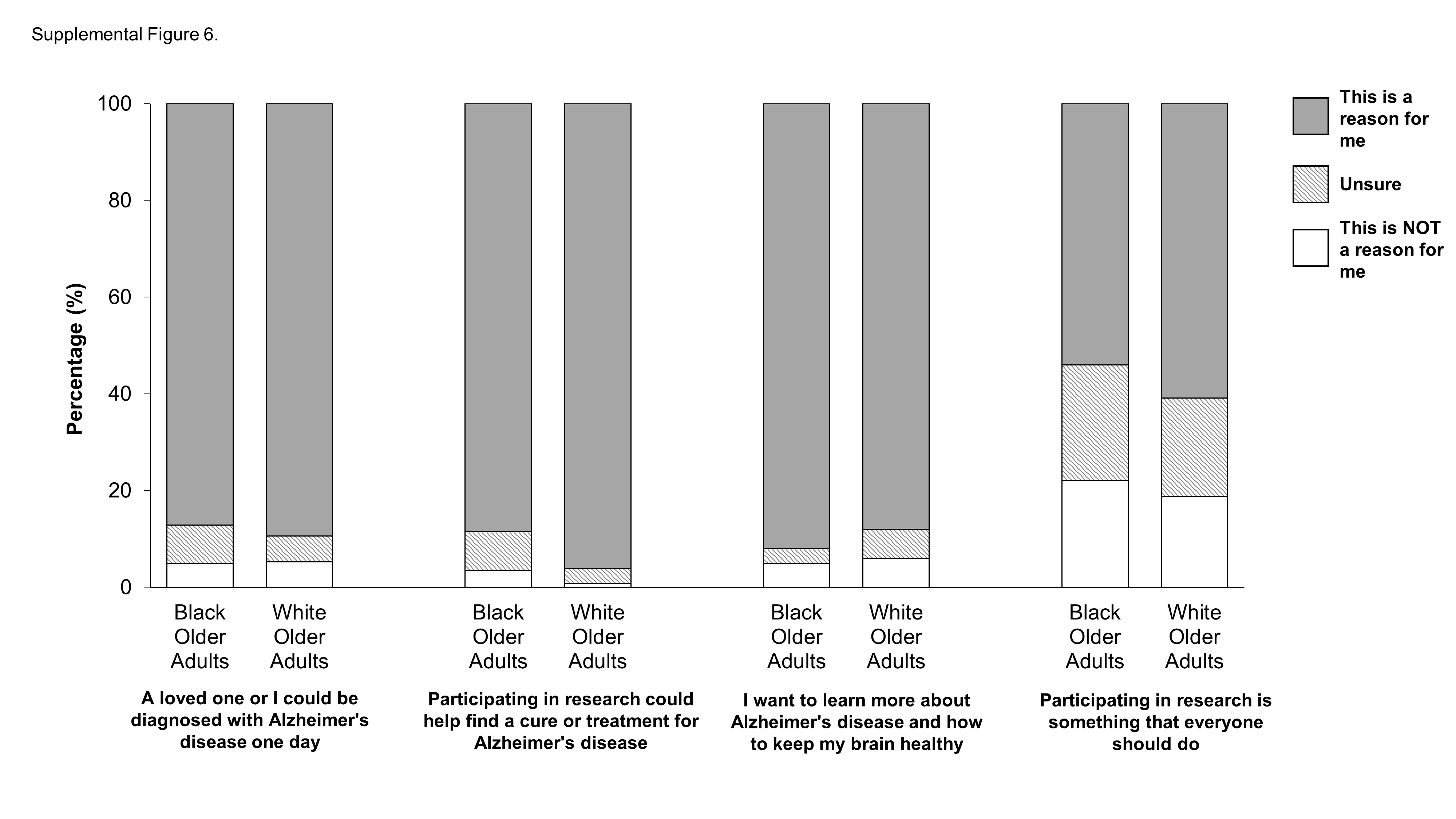
