## Supplemental Figure 2 for "Differences in Motivators, Barriers, and Incentives between Black and White Older Adults for Participation in Alzheimer’s Disease Biomarker Research"

**Questions below represent a subset of the questions asked on the survey that are relevant to the present study and manuscript.*

| **Questions** | **Responses** |
|---|---|
| Brain Health | |
| **1.** On a scale of 1 to 5, how would you rate how much you know about Alzheimer’s disease? | ○ I know nothing at all<br>○ I have heard of Alzheimer's disease, but I am not sure what it is<br>○ I know what Alzheimer's disease is<br>○ I know what Alzheimer's disease is and what causes it<br>○ I know what Alzheimer's disease is, what causes it, and how to manage and prevent it |
| Research Participation | |
| **2.** Have you ever been invited to participate in an Alzheimer's disease research study? | ○ No, no one ever asked me<br>○ Yes. I have been invited but DID NOT participate. |
| **2A**. In the future, if you were invited to participate in an Alzheimer's disease research study that involves giving blood to study genes for Alzheimer's disease and doing brain scans, would you consider doing it? | ○ No<br>○ Yes<br>○ Unsure |
| **3.** Which of the following statements are reasons why you may want to participate in Alzheimer’s disease research? | |
| **3A.** A loved one or I could be diagnosed with Alzheimer's disease one day. | ○ This is a reason for me<br>○ This is NOT a reason for me<br>○ Unsure |
| **3B.** Participating in research could help find a cure or treatment for Alzheimer's disease. | ○ This is a reason for me<br>○ This is NOT a reason for me<br>○ Unsure |
| **3C.** I want to learn more about Alzheimer's disease and how to keep my brain healthy. | ○ This is a reason for me<br>○ This is NOT a reason for me<br>○ Unsure |
| **3D.** Participating in research is something that everyone should do. | ○ This is a reason for me<br>○ This is NOT a reason for me<br>○ Unsure |

| | |
|---|---|
| **3E.** Other (please describe) | ○ This is a reason for me<br>○ This is NOT a reason for me<br>○ Unsure |
| **4.** Which of the following statements are reasons why you DID NOT participate in an Alzheimer's disease research study? | |
| **4A.** I was not qualified for the study. | ○ This is a reason for me<br>○ This is NOT a reason for me<br>○ Unsure |
| **4B.** I did not know much about Alzheimer's disease or the research process. | ○ This is a reason for me<br>○ This is NOT a reason for me<br>○ Unsure |
| **4C.** No one on the study represents people from my racial/ethnic background. | ○ This is a reason for me<br>○ This is NOT a reason for me<br>○ Unsure |
| **4D.** I was not worried about Alzheimer's disease affecting me. | ○ This is a reason for me<br>○ This is NOT a reason for me<br>○ Unsure |
| **4E.** The study was inconvenient (required too much time, travel, etc.) | ○ This is a reason for me<br>○ This is NOT a reason for me<br>○ Unsure |
| **4F.** I was afraid they would diagnose me with Alzheimer's disease. | ○ This is a reason for me<br>○ This is NOT a reason for me<br>○ Unsure |
| **4G.** I was concerned about how the study procedures would affect my health. | ○ This was a reason for me<br>○ This was NOT a reason for me<br>○ Unsure |
| **4H.** I did not trust the researchers. | ○ This was a reason for me<br>○ This was NOT a reason for me<br>○ Unsure |
| **4I.** I had bad experiences from previous research studies or medical procedures. | ○ This was a reason for me<br>○ This was NOT a reason for me<br>○ Unsure |
| **4J.** I would consider participating in a study now, but I need more information. | ○ This was a reason for me<br>○ This was NOT a reason for me<br>○ Unsure |
| **4K.** Other (please describe) | ○ This was a reason for me<br>○ This was NOT a reason for me<br>○ Unsure |

| **5.** Which of the following statements are reason(s) you WOULD NOT CONSIDER OR ARE UNSURE ABOUT participating in Alzheimer’s disease research? | |
|---|---|
| **5A.** I do not know enough about Alzheimer's disease to make a decision about participating in a research study. | ○ This is a reason for me<br>○ This is NOT a reason for me<br>○ Unsure |
| **5B.** I will not make any difference by participating in a research study. | ○ This is a reason for me<br>○ This is NOT a reason for me<br>○ Unsure |
| **5C.** I worry that the study staff/ researchers will use my information without my permission. | ○ This is a reason for me<br>○ This is NOT a reason for me<br>○ Unsure |
| **5E.** I worry about what they may find in my brain or test results. | ○ This is a reason for me<br>○ This is NOT a reason for me<br>○ Unsure |
| **5F.** I worry about the risks of the study. | ○ This is a reason for me<br>○ This is NOT a reason for me<br>○ Unsure |
| **5G.** I am not interested in Alzheimer's disease research. | ○ This is a reason for me<br>○ This is NOT a reason for me<br>○ Unsure |
| **5H.** I would consider participating in a study now, but I need more information. | ○ This is a reason for me<br>○ This is NOT a reason for me<br>○ Unsure |
| **5I.** Other (please describe) | ○ This is a reason for me<br>○ This is NOT a reason for me<br>○ Unsure |
| **6.** Please select your opinion about each of these statements. Please note the answer in the left column starts with disagree. | |
| I trust researchers to keep my information confidential. | ○ Disagree<br>○ Agree<br>○ Not sure/Do Not Know |
| I trust researchers to be honest with me. | ○ Disagree<br>○ Agree<br>○ Not sure/Do Not Know |
| I trust researchers to be competent. | ○ Disagree<br>○ Agree<br>○ Not sure/Do Not Know |
| I trust researchers will not harm me. | ○ Disagree<br>○ Agree<br>○ Not sure/Do Not Know |

| | |
|---|---|
| I trust researchers to treat me equally, with acceptance and respect regardless of my age, gender, race/ethnicity, sexual orientation, etc. | ○ Disagree<br>○ Agree<br>○ Not sure/Do Not Know |
| I trust researchers to share what they learned with me and my community. | ○ Disagree<br>○ Agree<br>○ Not sure/Do Not Know |
| Other (please describe) | ○ Disagree<br>○ Agree<br>○ Not sure/Do Not Know |
| Perceptions about Research Procedures | |
| The next set of questions ask about whether you would consider doing certain medical procedures as part of a research study. Please note that we are NOT asking you to do any procedures as part of this survey. | |
| In the future, if I were to be invited to participate in an Alzheimer’s disease research study... | |
| **8.** I would consider giving a blood sample to test for genes which may affect the risk of getting Alzheimer's disease. | ○ No<br>○ Yes<br>○ Maybe in the future |
| **9.** Which of the following statements best describe why you would not give a blood sample in an Alzheimer's disease research study? | |
| **9A.** I do not have enough information about this procedure to make a decision. | ○ This is a reason for me<br>○ This is NOT a reason for me<br>○ Unsure |
| **9B.** I do not think this procedure is safe. | ○ This is a reason for me<br>○ This is NOT a reason for me<br>○ Unsure |
| **9C.** I do not know the doctors or staff who will do the procedure. | ○ This is a reason for me<br>○ This is NOT a reason for me<br>○ Unsure |
| **9D.** I do not know what will happen to my information. | ○ This is a reason for me<br>○ This is NOT a reason for me<br>○ Unsure |
| **9E.** I only do these procedures when they are medically necessary. | ○ This is a reason for me<br>○ This is NOT a reason for me<br>○ Unsure |
| **9F.** I am worried about abnormal test results. | ○ This is a reason for me<br>○ This is NOT a reason for me<br>○ Unsure |

| | |
|---|---|
| **10.** I would consider doing a brain MRI for Alzheimer’s disease research.<br>[The MRI shows pictures of the brain structure.] | ○ No<br>○ Yes<br>○ Maybe in the future |
| **11.** Which of the following statements best describe why you would not get a brain MRI in an Alzheimer's disease research study? | |
| **11A.** I do not have enough information about this procedure to make a decision. | ○ This is a reason for me<br>○ This is NOT a reason for me<br>○ Unsure |
| **11B.** I do not think this procedure is safe. | ○ This is a reason for me<br>○ This is NOT a reason for me<br>○ Unsure |
| **11C.** I do not know the doctors or staff who will do the procedure. | ○ This is a reason for me<br>○ This is NOT a reason for me<br>○ Unsure |
| **11D.** I do not know what will happen to my information. | ○ This is a reason for me<br>○ This is NOT a reason for me<br>○ Unsure |
| **11E.** I only do these procedures when they are medically necessary. | ○ This is a reason for me<br>○ This is NOT a reason for me<br>○ Unsure |
| **11F.** I am worried about abnormal test results. | ○ This is a reason for me<br>○ This is NOT a reason for me<br>○ Unsure |
| **12.** I would consider doing a PET scan for Alzheimer’s disease research.<br>[The PET scan is similar to a MRI, but it can detect brain damage from Alzheimer’s disease. It requires getting an injection with a dye, which leaves your system after a few hours.] | ○ No<br>○ Yes<br>○ Maybe in the future |
| **13.** Which of the following statements best describe why you would not get a PET scan in an Alzheimer's disease research study? | |
| **13A.** I do not have enough information about this procedure to make a decision. | ○ This is a reason for me<br>○ This is NOT a reason for me<br>○ Unsure |
| **13B.** I do not think this procedure is safe. | ○ This is a reason for me<br>○ This is NOT a reason for me<br>○ Unsure |
| **13C.** I do not know the doctors or staff who will do the procedure. | ○ This is a reason for me<br>○ This is NOT a reason for me<br>○ Unsure |

| | |
|---|---|
| **13D.** I do not know what will happen to my information. | ○ This is a reason for me<br>○ This is NOT a reason for me<br>○ Unsure |
| **13E.** I only do these procedures when they are medically necessary. | ○ This is a reason for me<br>○ This is NOT a reason for me<br>○ Unsure |
| **13F.** I am worried about abnormal test results. | ○ This is a reason for me<br>○ This is NOT a reason for me<br>○ Unsure |
| Incentives | |
| **14.** If you were to participate in an Alzheimer's disease research study and had to do blood draw, brain MRI, and PET scans, what incentives would you like to receive in addition to a gift card?<br><br>Check all that apply. | ○ Results of routine blood work (such as glucose and cholesterol tests)<br>○ Results of brain scans<br>○ Results of memory tests<br>○ Transportation vouchers or free rideshares such as Uber or Lyft<br>○ Information about brain health<br>○ Does not apply. I would not do any of these procedures<br>○ Other |
| **15.** Should normal results from the blood test and brain MRI be shared with the participant? (NOTE: Abnormal test results from these procedures are already being shared with participants.) | ○ No<br>○ Yes<br>○ Unsure |
