## Supplementary material for "Differences in Motivators, Barriers, and Incentives between Black and White Older Adults for Participation in Alzheimer’s Disease Biomarker Research": List of supplemental Figures

**Supplemental Figure 1**. Diagram outlining contacted, eligible, and ineligible participants from the community, VA, and PowerTrials.

**Supplemental Figure 2.** Select questions from the AD-REACH survey relevant to the present study.

**Supplemental Figure 3.** (A) Flow of the overall sets of questions in the survey; (B) survey question flow for assessing interest in Alzheimer’s disease (AD) research and motivators and barriers to participation in AD research in general; and (C) survey question flow for interest in and barriers to specific AD biomarker procedures (blood draw, MRI, PET).

**Supplemental Figure 4.** Reasons for not enrolling in Alzheimer’s disease research despite previously being asked to participate (n = 14)

**Supplemental Figure 5.** Percentage of Black and White participants endorsing barriers to participating in (A) general Alzheimer’s disease biomarker research (n = 118), (B) blood draw for genetics (n = 89), (C) MRI (n = 115), and (D) PET (n = 138).

**Supplemental Figure 6**. Percentage of Black and White participants endorsing different motivations for participating in general Alzheimer’s disease biomarker research (n = 359).
