## Supplemental Table 1 for "Differences in Motivators, Barriers, and Incentives between Black and White Older Adults for Participation in Alzheimer’s Disease Biomarker Research"

**Supplementary Table 1**. Association between incentives and interest in participating in an AD biomarker study (n = 399).

| | 95% CI | | | | |
|---|---|---|---|---|---|
| | OR | lower | upper | *p* | OR for interest to participate in study |
| Results of routine blood work | 0.25 | 0.15 | 0.42 | < .001 | 4.02 |
| Results of brain scans | 0.22 | 0.12 | 0.40 | **< .001** | 4.50 |
| Results of memory tests | 0.28 | 0.16 | 0.50 | **< .001** | 3.58 |
| Transportation vouchers or free rideshares | 0.67 | 0.40 | 1.01 | **.120** | 1.50 |
| Information about brain health | 0.44 | 0.26 | 0.72 | **.001** | 2.29 |

*NOTE: OR = odds ratio; CI = confidence interval. Cumulative OR and 95% CI were calculated using ordinal logistic regression models to examine the association between interest in an incentive and hesitancy to participate in an AD biomarker study. OR for interest to participate in an AD biomarker study was calculating by taking the inverse of the cumulative OR. Questions about monetary incentives were not included in this survey.*
